# Lesions causing Alice in Wonderland Syndrome map to a common brain network linking body and size perception

**DOI:** 10.1101/2024.01.17.24301332

**Authors:** Maximilian U. Friedrich, Elijah C. Baughan, Isaiah Kletenik, Ellen Younger, Charlie W. Zhao, Calvin Howard, Michael A. Ferguson, Amalie Chen, Daniel Zeller, Claudia Piervincenzi, Silvia Tommasin, Patrizia Pantano, Olaf Blanke, Sashank Prasad, Jared A. Nielsen, Michael D. Fox

## Abstract

**Background:** In Lewis Carroll’s 1865 novel “Alice’s Adventures in Wonderland”, the protagonist experiences distortions in the size of her body and those of others. This fiction becomes reality in neurological patients with Alice in Wonderland Syndrome (AIWS). Brain lesions causing AIWS may offer unique insights into the syndrome’s elusive neuroanatomy.

**Methods:** A systematic literature search identified 37 cases of lesion-induced AIWS. Lesion locations were mapped onto a brain atlas and functional connectivity between each lesion location and other brain regions was estimated using resting-state fMRI data from 1000 healthy subjects. Connections common to AIWS lesions were identified and compared to connections from 1073 lesions associated with 25 other neuropsychiatric disorders. Alignment between this lesion-derived AIWS network and neuroimaging findings from patients with AIWS due to other etiologies was assessed.

**Results:** Although AIWS lesions occurred in many different brain locations, these lesions fell within a specific, functionally connected brain network. This network was defined by connectivity to the right extrastriate body area, a brain region selectively activated by viewing body parts, and the inferior parietal cortex, a brain region involved in processing of size and scale. This connectivity pattern was specific to AIWS when compared to lesions causing other neuropsychiatric disorders and aligned with neuroimaging findings in patients with AIWS from other etiologies.

**Conclusion:** Lesions causing AIWS fall within a specific brain network defined by connectivity to two distinct brain regions, one region involved in body perception and another in processing of size and scale.

## Introduction

In Lewis Carroll’s 1865 classic novel “Alice’s Adventures in Wonderland”, the protagonist experiences a surreal world with curiously warped rules of size and space. Most notably, these distortions affect the perception of her own body or those of the Wonderland’s inhabitants (Figure 1A, C). Similar distortions have been reported by patients with neurological disorders. Early reports include a patient with migraine who reported “my neck stretches and my head goes to the ceiling”^1^. In fact, Lewis Carroll himself may have had migraine, raising the possibility that his fiction mirrored personal experience^2,3^. The striking parallels between Alice’s experiences and these symptoms led to the coining of the term of Alice in Wonderland Syndrome (AIWS) in 1955^4^.

**Figure 1.**
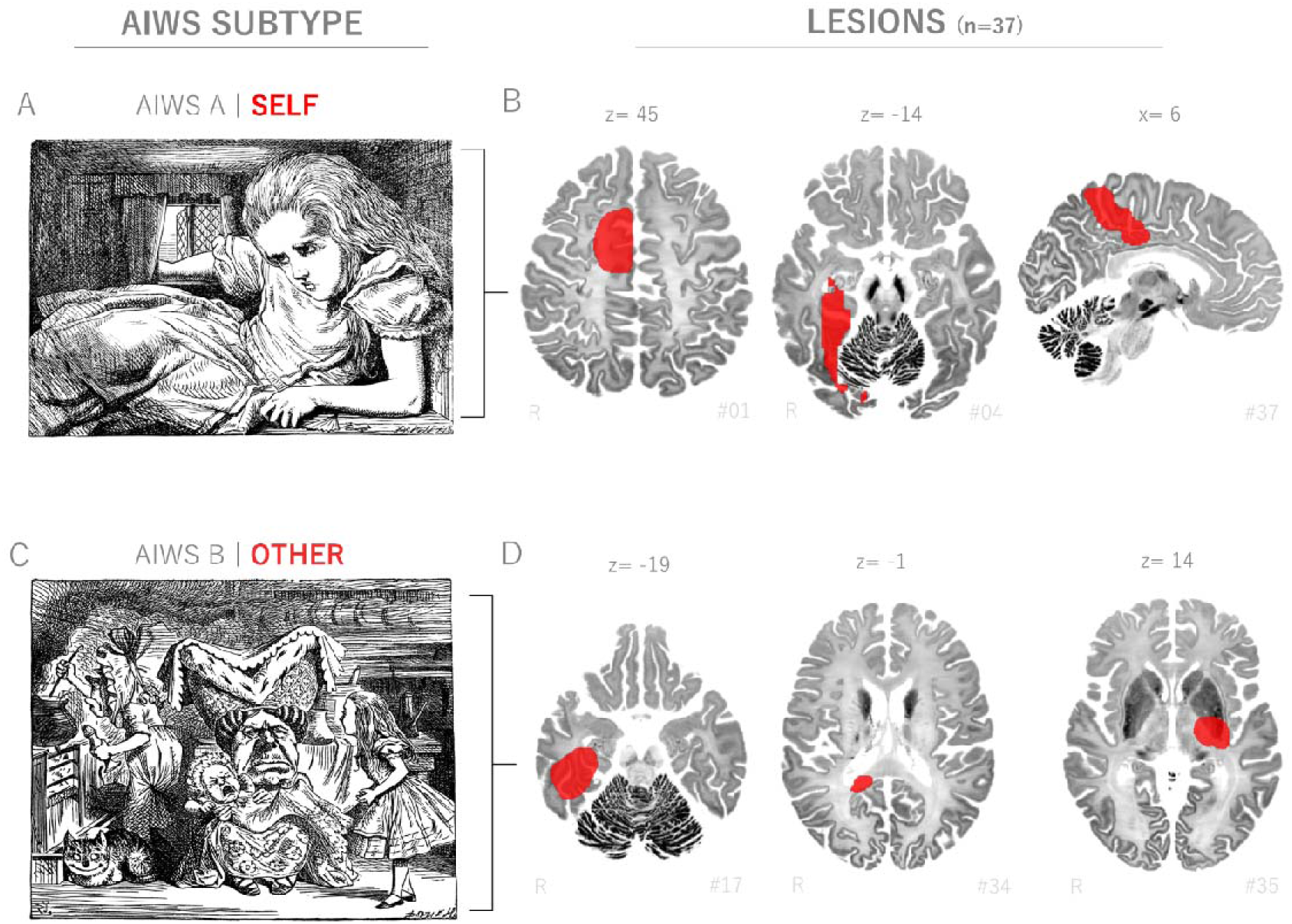
Lesions causing AIWS are neuroanatomically and clinically heterogeneous. A) Artistic illustration shows the protagonist Alice perceiving enlargement of her own body, including disproportionate enlargement of her head. B) Lesion locations associated with AIWS-Self, i. e. perceptual distortions related to one’s own body (red, showing 3/6 cases). C) Artistic illustration shows Alice observing other figures whose body proportions are remarkably altered. D) Lesion locations associated with AIWS-Other (red, showing 3 of 29 cases), i. e. perceptual distortions related to bodies and objects in surrounding space. Lesions depicted in radiological convention (R= right). Original illustrations by Sir John Tenniel (1865), public domain, distributed via project Gutenberg^35^.

Since that time, around 170 cases of AIWS have been described in the medical literature and it was found that up to 38% of healthy adolescents report transient AIWS-like symptoms (reviewed in ^3,5^). Most frequently, patients perceive their own body or others’ bodies/ objects in space as too large or too small^3,5^, which has led to the proposal of different clinical subtypes^3,5^.

Despite a long-standing scientific interest, the pathophysiology and neuroanatomy of AIWS remains unclear. AIWS has variably been classified as a hallucination^1^, an illusory misperception^3,4^, or a disorder of multisensory integration related to body representation^3,5–8^. Functional neuroimaging studies in patients experiencing AIWS have noted abnormalities, but these have been reported in multiple different brain regions^9–13^. Migraine is the most common cause of AIWS (∼27%). The next most common cause is focal brain damage (∼8%)^5^, which can allow for causal links to human neuroanatomy^14^. However, lesions causing AIWS have been reported in multiple different brain locations with little to no overlap^15^. It remains unclear how damage to a single brain region, and to different regions in different patients, can lead to the complex, yet stereotypical perceptual distortions characteristic of AIWS.

Lesion network mapping is a recently validated method that was designed to help address these questions^16,17^. By examining the connections between lesion locations and other brain regions that may be distant from the lesion location itself, one can gain insight into how lesions in different locations can cause the same syndrome^18–20^ and how a single lesion can lead to simultaneous disruptions in multiple different brain functions^19,21,22^.

Here, we utilize lesion network mapping to test the hypothesis that lesion locations causing AIWS fall within a common brain network, defined by connectivity to a specific set of distant brain regions.

## Methods

See supplementary information for additional details.

### Research ethics

This retrospective study was approved by the institutional review board of Brigham & Women’s Hospital, Boston, MA, USA (#2020P002987).

### Case collection

Reported cases of lesion-induced AIWS were collated from two independently conducted systematic literature searches following the “Preferred Reporting Items for Systematic reviews and Meta-Analyses” (PRISMA) guidelines^23^ (see supplementary information for details). The initial search, concluding in November 2021, identified 30 cases from 26 studies which have been previously documented^15^. The subsequent search, concluding in September 2023, identified an additional 9 cases of lesion-induced AIWS.

### Inclusion criteria

We identified cases that met three criteria: 1) at least one of the syndrome-defining perceptual distortions, 2) presence of a focal brain lesion temporally associated with symptom expression, 3) at least one documentation of neuroimaging or neuropathology of sufficient quality for precise lesion identification and delineation. Applying these criteria led to exclusion of two cases from the initial search, resulting in a total of 37 cases for our main analysis (Supplementary Table 1). To ensure that our results were not dependent on our specific inclusion criteria or our new secondary search, we repeated the main analyses using the full 30 cases identified in the initial search^15^.

### Lesion network mapping

We used a technique called “lesion network mapping” to understand the connections between each lesion and the rest of the brain^16,17^. This method involves the following steps:

### Lesion segmentation

As in prior studies^16,17^, lesion locations were copied by hand from figures appearing in the original publications onto a reference brain atlas in standard space (Montreal Neurological Institute 152, MNI152, 2mm isotropic) using MRIcroGL (https://www.nitrc.org/projects/mricrogl). To ensure reproducibility, two independent raters (MUF, CWZ) mapped all lesions, consistent with current best practice recommendations. In cases where published images displayed multiple neuroimaging slices (54% of reports), all slices were copied in the corresponding axis, providing a more robust approximation of full lesion extent. Previous work shows that the resulting 2D lesion traces are suitable for network mapping^18^.

### Lesion network overlap

Next, functional connectivity between each lesion location and all other brain voxels was determined using resting-state functional MRI (rsfMRI) data from 1000 healthy, adult subjects from the “genome superstruct project” (50% females)^24,25^. RsfMRI measures spontaneous fluctuations in brain activity by tracking changes in blood oxygenation. Regions with correlated activity are considered functionally connected. By combining results across 1000 individuals, we obtain a robust estimate of brain regions functionally connected to each lesion location. Each voxel in the resulting lesion network map is assigned a T-value as a statistical measure of how strongly the lesion location is connected to that voxel. Positive T-values signify a positive correlation and a synchronized pattern of activity, while negative T-values indicate an anticorrelation^26^ and an opposite pattern of activity.

To assess the consistency of lesion connection in the AIWS cohort, we thresholded the lesion network maps at |T|≥ 5 (voxel-wise family-wise error (FWE)-corrected p< .05), creating binarized maps of brain regions significantly connected to the lesion location. We repeated this process at |T|≥ 7 (voxel-wise FWE-p< 10^-6^) and |T|≥ 9 (voxel-wise FWE-p< 10^-11^) to verify robustness against threshold variations. Finally, we overlaid the binarized lesion network maps to identify regions with consistently shared connections across the multiple different lesions in the AIWS cohort. This approach reflects the current standard of the lesion network mapping methodology^16^.

### Specificity testing

To assess specificity, we contrasted the unthresholded lesion network maps from the AIWS lesions with 1073 lesion network maps derived from lesions causing 25 other neuropsychiatric disorders previously reported by our group, including several other perceptual disorders (e. g. hallucinations^19^, prosopagnosia^21^, central visual and vestibular disorders^18,27^). A two-sample t-test, accounting for lesion size as a nuisance variable, was implemented using publicly available software for statistical analysis of neuroimaging data (FSL’s permutation analysis of linear models, PALM^28^). To test for connections that might be specific to one AIWS subtype, we performed a similar two-sample t-test comparing lesion networks from AIWS-Self to AIWS-Other. Results were corrected for multiple comparisons using voxel-based family-wise error correction in FSL PALM.

### Deriving a lesion-based AIWS network

A conjunction map was created from the lesion network overlap map and the specificity map to identify brain regions with consistent (i. e. connected in ≥85% of cases) and specific (i. e. regions with FWE-p< .05 in specificity analysis) connectivity to AIWS lesions. We will refer to the areas resulting from this conjunction analysis as “hubs” of the AIWS network. We compared the location of these hubs to standard atlases of brain structure and function including the probabilistic functional atlas of the human occipito-temporal visual cortex^29^, the Juelich probabilistic cytoarchitectonic atlas^30^ and the brainnetome atlas^31^. Hub locations and atlas boundaries were compared by overlaying both on a high-resolution MRI template (400µm isotropic, Juelich EBrains) in FSLeyes^32^ or MRIcroGL. Using these hubs, we also conducted a post-hoc analysis to estimate the effect size (Cohen’s d) of our connectivity findings. We computed each lesion’s connectivity to the network hubs and compared AIWS lesions to those associated with other neuropsychiatric disorders using a two-sample t-test.

By definition, positive connectivity to the positive AIWS hub and negative connectivity to the negative AIWS hub defines a distributed brain network best encompassing the lesion locations associated with AIWS, while avoiding control lesions associated with other syndromes. To visualize this network, we computed functional connectivity with the two AIWS network hubs, thresholded each connectivity map at |T|= 7 and identified all voxels that satisfied the above criteria. We also computed a “sign inverted” AIWS network with the opposite connectivity patterns (i. e. negative connectivity to positive network hub and vice versa). This hypothetical network encompasses brain locations that, when hyperactivated or stimulated, might theoretically cause AIWS.

### Relating AIWS lesions to other AIWS etiologies

We focused first on migraine^33,34^ because it is the most common AIWS etiology^3,5^ and because a migraine network has recently been defined by connectivity to a hub area in the left extrastriate cortex (area V3)^33^. We computed the connectivity of area V3^29,33^, calculated the Pearson correlation to lesion network maps associated with AIWS and compared it to those associated with other neuropsychiatric disorders. Absolute Pearson r-values reflect the degree of similarity, while the sign indicates the direction of relationship (positive values indicate similar, negative values opposite connectivity patterns).

To test alignment of the lesion-derived AIWS network with functional neuroimaging studies of AIWS due to non-lesion etiologies, an additional systematic literature search was carried out to identify relevant studies (see supplementary information). Five studies included sufficient documentation of functional neuroimaging (i. e. single-photon emission CT (SPECT), functional MRI) alterations during AIWS episodes to transfer the reported locations into reference space. Overlap was determined between hypoactivation locations and the lesion-derived AIWS network as well as hyperactivation locations and the negative, or “sign-inverted” AIWS network. Finally, locations of hypoactivation or hyperactivation were combined into two separate ROIs, their connectivity was computed and the resulting network maps were correlated to those associated with AIWS and other neuropsychiatric disorders for comparison.

### Statistical computations

Statistical computations were conducted with Jamovi Version 2.4 (www.jamovi.org), GraphPad Prism Version 9 (www.graphpad.com) and Python Version 3 (www.python.org) with Pandas, NumPy, Nibabel, seaborn, SciPy packages. Data distributions were determined using Shapiro-Wilk testing and inspection of interquartile (“Q-Q- “) plots. To contrast group means, parametric tests with appropriate control of unequal variances (e. g. two-sample t-test with Welch’s correction after Levene’s test) were used, unless the assumption of normality was violated. In this case, non-parametric alternatives were used (e. g. Wilcoxon-Mann-Whitney test). Results from voxel-wise analyses were corrected for multiplicity using the family-wise error method in FSL PALM. Data is reported as mean ± standard deviation or median and interquartile range. The significance level was set at p< .05 (flagged as *, p< .01 as ** and p< .001 as ***). Alongside p-values, 95% confidence intervals of the mean of differences are reported as a measure of effect size.

## Results

### Clinical characteristics

Our systematic literature search identified 37 AIWS cases caused by focal brain damage: 16.2% (6/37) involved perceptual distortions relating to the patients’ own bodies (AIWS-Self), 78.4% (29/37) involved perceptual distortions relating to other’s bodies or objects in space (AIWS-Other), and the remaining 5.4% (2/37) involved overlapping features (AIWS-Mixed). Most AIWS-Other cases involved visual distortion of bodies or body parts (87%), while most AIWS-Self cases involved somatosensory distortion of one’s own body (75%).

Across all subtypes, perceptual distortions most frequently involved the head or face (38%, 14/37), followed by the upper extremity or hand (24%, 9/37). In 54% of cases, more than one perceptual distortion was reported, including exaggerated movement (kinetopsia, 3/37), difficulty discriminating colors (dyschromatopsia, 2/37), and persistent after-images (palinopsia, 1/37). Detailed cohort characteristics are shown in Table 1 and Table S2.

**Table 1.** Clinical characteristics of AIWS cases.

|  | AIWS-All | AIWS-Self | AIWS-Other | AIWS-Mixed |
| --- | --- | --- | --- | --- |
| **N** | 37 | 6 | 29 | 2 |
| **Age (y)** | 62.1 ± 14.6 | 53.7 ± 7.2 | 63.1 ± 15.3 | 73.5 ± 13.4 |
| **Gender** | 21M / 16W | 5M / 1W | 14M / 15W | 2M / 0W |
| **Cortical visual deficit (%)** | 40.5 | 16.7 | 48.3 | 0.0 |
| **Neglect (%)** | 16.2 | 0.0 | 21.0 | 0.0 |
| **Anosognosia (%)** | 11.0 | 16.7 | 10.3 | 0.0 |
| **Macropsia (%)** | 41.0 | 0.0 | 48.3 | 50.0 |
| **Micropsia (%)** | 62.0 | 0.0 | 72.4 | 100 |
| **Telopsia (%)** | 11.0 | 0 | 14.0 | 0 |
| **Pelopsia (%)** | 11.0 | 0 | 14.0 | 0 |
| **Macrosomatognosia (%)** | 13.5 | 67.0 | 0.0 | 50.0 |
| **Microsomatognosia (%)** | 9.8 | 33.0 | 0.0 | 100 |
| **Visual distortions primarily body-related (%)** | NA | NA | 65.0 | 100 |
| **Visual distortions body- and object-related** | NA | NA | 87.0 | 0 |
| **Positive migraine history (%)** | 11.0 | 0.0 | 13.8 | 0.0 |

### Lesion topography

The majority of lesions causing AIWS affected supratentorial structures (92%, 34/37) and were located in the right hemisphere (70%, 26/37, binomial proportion test: p= .02). Lesion laterality did not significantly differ between AIWS subtypes (Fisher’s exact test, p= .15). Lesion locations were highly heterogeneous, with less than a third of lesions affecting the same brain region (Figure 1). The most commonly lesioned brain region was the right extrastriate occipital cortex followed by the right temporal cortex (Supplementary Table 2).

### Lesions causing AIWS map to a common brain network

Despite this heterogeneity in lesion location and clinical presentation, lesion locations causing AIWS were functionally connected to a common set of brain regions (Figure 2). Specifically, over 90% of lesion locations were positively connected to a circumscribed area in the right lateral occipital cortex and over 85% were negatively connected to a region in the left parietal cortex. This connectivity profile was consistent across AIWS subtypes (Figure 2 and Supplementary Figure 2). Results were similar when different individuals traced the lesion locations (mean Pearson r= 0.96 ± 0.11, Supplementary Figure 3), when different statistical thresholds were applied (Pearson r= 0.95 – 0.98, Supplementary Figure 4) and when different systematic search criteria were used (Pearson r= 0.76 – 0.89, Supplementary Figure 5).

**Figure 2.**
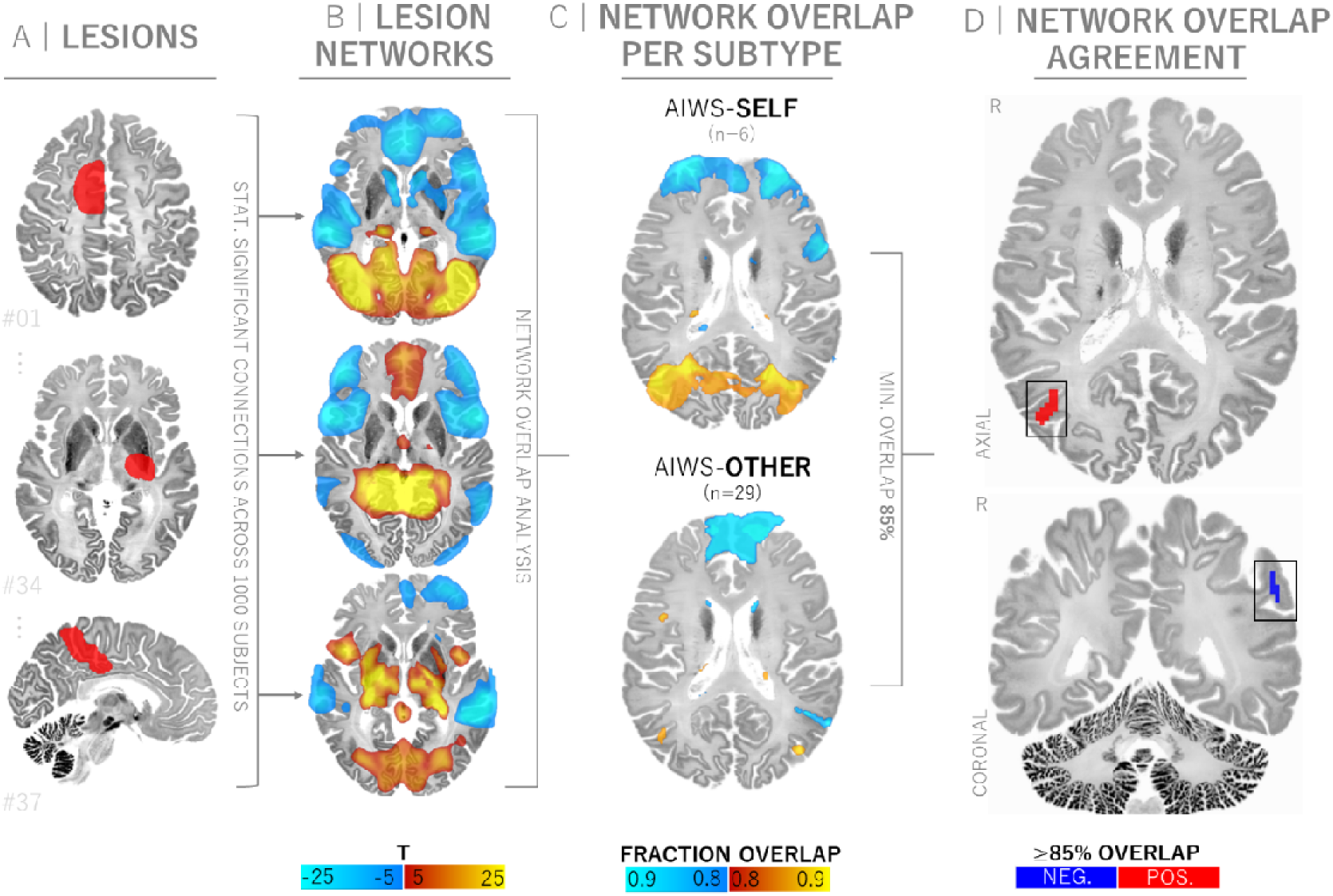
Lesions causing AIWS map to a common brain network. A) Each individual lesion location (red) associated with AIWS was mapped to a common brain atlas. B) Functional connectivity between each lesion location and all other brain voxels was computed using a database of brain connectivity derived from 1000 healthy subjects, generating a lesion network for each lesion location. C) Lesion networks from individual cases were overlapped to identify brain regions connected to all or most lesion locations. Separate overlap analyses for the AIWS-Self (6 cases) and AIWS-Other (29 cases) subtypes were conducted. D) Brain regions connected to > 85% of AIWS lesion locations, irrespective of AIWS subtype. Positive connectivity is depicted in warm colors and red, negative connectivity (or anticorrelation) in cool colors and blue.

The observed connectivity pattern was specific to lesions causing AIWS compared to 1073 lesions associated with 25 other neuropsychiatric disorders (Figure 3, voxel-wise FWE-p< .05). A similar analysis contrasting lesion networks from different AIWS subtypes yielded no significant differences. A conjunction analysis identified two clusters both sensitive and specific to lesions causing AIWS: a positively connected region in the right occipital cortex (centered at X= 42, Y= 71, Z= 16 MNI, cluster volume 1350mm^3^, d= 1.2, 95% CI [0.87, 1.53]) and a negatively connected region in the left inferior parietal cortex (−57, −51, 37, 900mm^3^, d= −0.97, 95% CI [-1.30, - 0.64], Figure 3).

**Figure 3.**
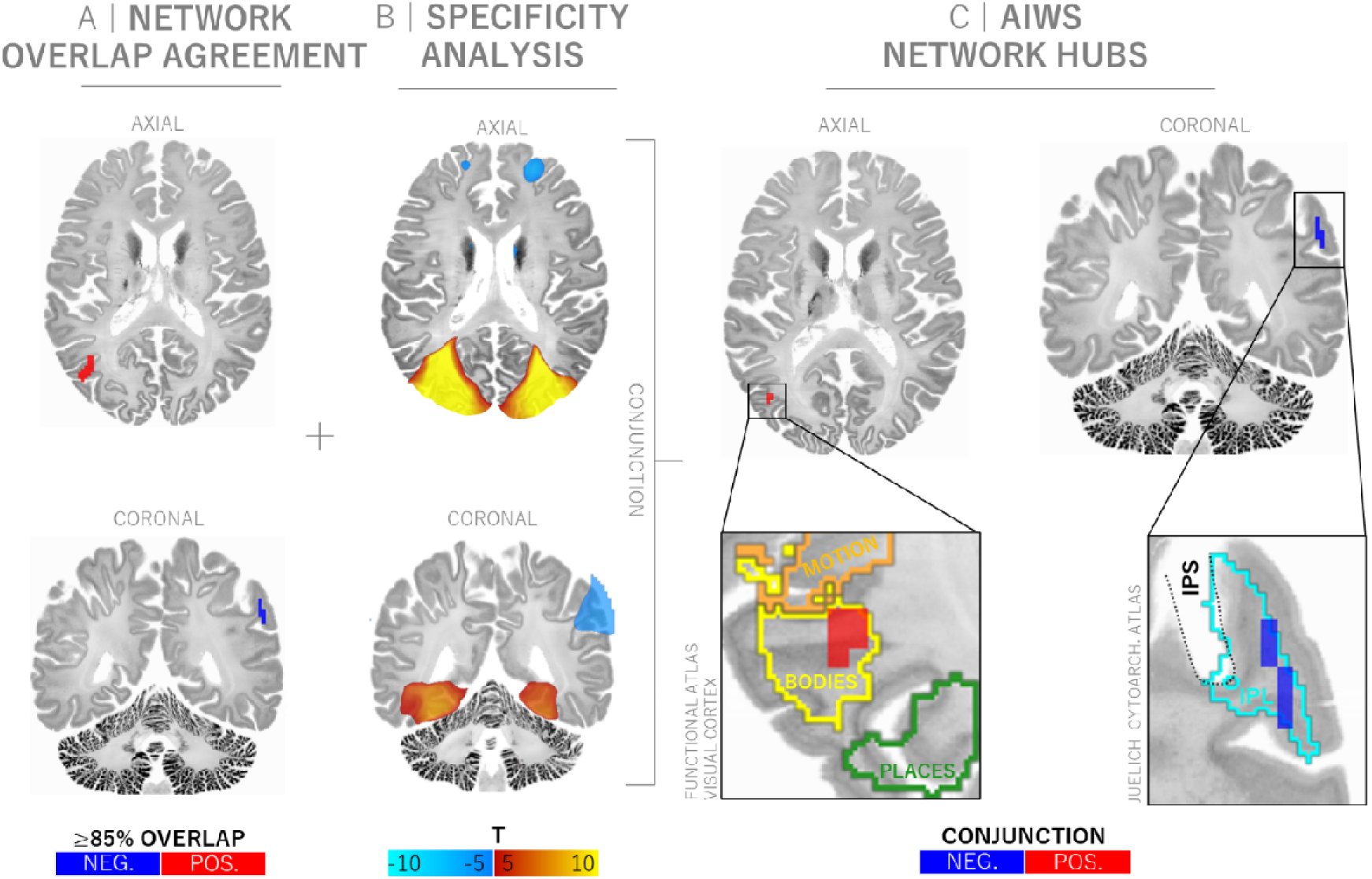
AIWS network hubs align with brain regions previously implicated in body perception and size processing. A) Network overlap agreement map (reproduced from figure 2D) showing regions connected to ≥85% of AIWS lesion locations. B) Specificity maps showing connections that are specific to AIWS lesion locations compared to 1073 lesion locations associated with other symptoms or syndromes (voxel-wise FWE-corrected p< .05). C) AIWS Network hubs are defined by connections that are both sensitive and specific to AIWS. The right occipital hub (red) aligns with the location of the extrastriate body area (yellow outline) which selectively responds to images of bodies or body parts. The left parietal hub (blue) aligns with the inferior parietal lobule (IPL, cyan outline) adjacent to the intraparietal sulcus’ (IPS, dashed black outline) implicated in quantitative processing including size estimation.

We will refer to these clusters as network hubs. Results were consistent when repeated for AIWS subtypes separately (Supplementary Figure 6). This conjunction analysis also yielded some secondary, smaller clusters (<200mm^3^, Supplementary Figure 7).

AIWS network hubs map to higher-order areas involved in body perception and quantity processing To contextualize the data-driven connectivity findings, we projected the identified AIWS network hubs onto MRI atlases that correlate function with anatomical parcellations. The occipital hub closely aligned with the right extrastriate body area (EBA), a higher-order visual area predominantly activated in response to seeing bodies and body parts^29,36–38^(Figure 3). The negatively connected hub was located in the left inferior parietal lobule (IPL), bordering the intraparietal sulcus (IPS1)^30^. This higher-order area has been implicated in quantity processing, including processing of sizes, magnitudes, distances and scales^39–42^, as well as mental rotation and social cognition^31,39^.

### The AIWS network encompasses neuroanatomically diverse AIWS lesion locations

By definition, positive connectivity to the right EBA hub and negative connectivity to the left IPL/IPS hub delineates the network of brain regions that, when lesioned, can cause AIWS. To visualize this network, we computed connectivity with these two regions (Figure 3C) and identified all brain voxels that met the above criteria. As expected, this network encompasses 97% (36/37) of the neuroanatomically heterogeneous lesion locations causing AIWS (Figure 4D). The only lesion that fell outside this network was located in the medulla (case 06).

**Figure 4.**
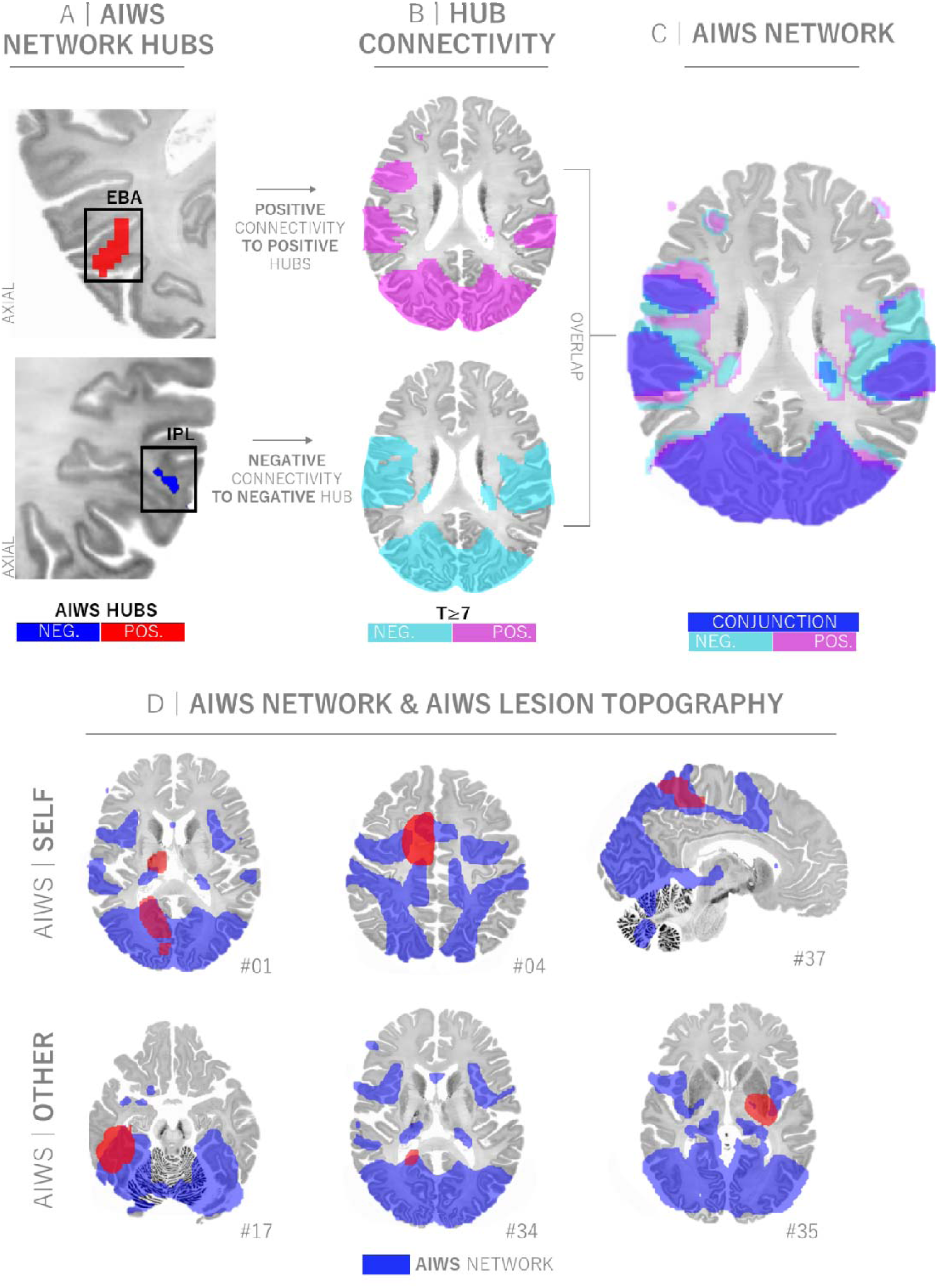
A single common brain network encompasses diverse lesion locations associated with AIWS. A) AIWS network hubs in the occipital cortex (red) and parietal cortex (blue). B) Functional connectivity with network hubs identifies voxels positively correlated with the occipital hub (pink) and negatively correlated with the parietal hub (cyan). C) Conjunction analysis identifies an AIWS network composed of voxels that meet both criteria (blue). D) As expected, this common AIWS network (blue) encompasses 97% (36/37) of the different lesion locations associated with AIWS (red). Exemplary lesion locations are identical to those shown in Figure 1.

### Alignment of AIWS lesions with other AIWS etiologies

Given that migraine is the most frequent cause of AIWS in adults^3,5^, we explored whether our lesion-derived network findings might align with neuroimaging findings in migraine. Although the neuroanatomy of migraine remains debated, multiple studies point to a pivotal role of extrastriate visual area V3 in the genesis of cortical spreading depression and migraine aura (Figure 5A)^33,34,43^. Like AIWS lesions, area V3 fell within the AIWS lesion network (Figure 5B). The connectivity of area V3 was more similar to the connectivity of AIWS lesions than to lesions associated with other neuropsychiatric disorders (median Pearson r= 0.75, p< .001, 95% CI [0.38, 0.79], Figure 5C).

**Figure 5.**
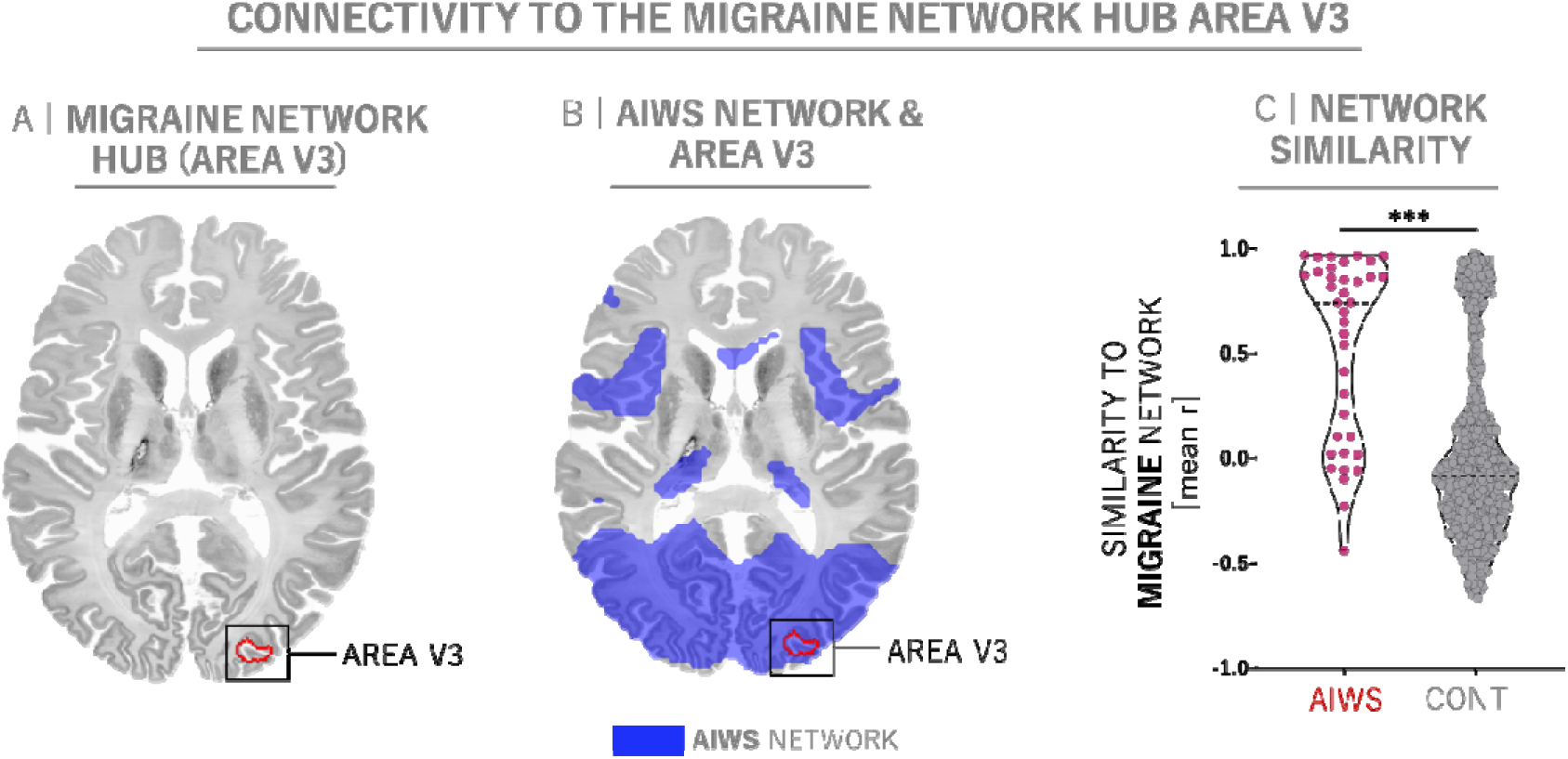
The lesion-derived AIWS network aligns with the migraine network. A) Area V3 (red outline), has been implicated in migraine aura and cortical spreading depression and B), falls within the boundaries of the lesion-based AIWS network (blue, reproduced from Figure 4). C) The connectivity of area V3 was more similar to the connectivity of AIWS lesions than to lesions associated with other neuropsychiatric disorders as controls (CONT, median Pearson r= 0.75, p< .001, 95% CI [0.38, 0.79]).

Next, we explored whether our results might align with functional neuroimaging findings from cases of AIWS caused by non-lesional etiologies. Across five cases, we found that locations of functional hypoactivation fell within the AIWS lesion network (Figure 6A). The connectivity patterns of AIWS-related hypoactivation were significantly more similar to lesions associated with AIWS than lesions associated other neuropsychiatric disorders (p< .001, 95% CI [0.14, 0.44], Figure 6B). In two of the five neuroimaging cases, additional regions of functional hyperactivation during AIWS episodes were reported. These foci aligned with the opposite pattern of the lesion-derived AIWS network (Figure 6C). AIWS-related hyperactivations showed an inverse connectivity pattern and were more negatively correlated to lesion networks associated with AIWS than other neuropsychiatric disorders (p< .001, 95% CI [-0.11, 0.35], Figure 6D).

**Figure 6.**
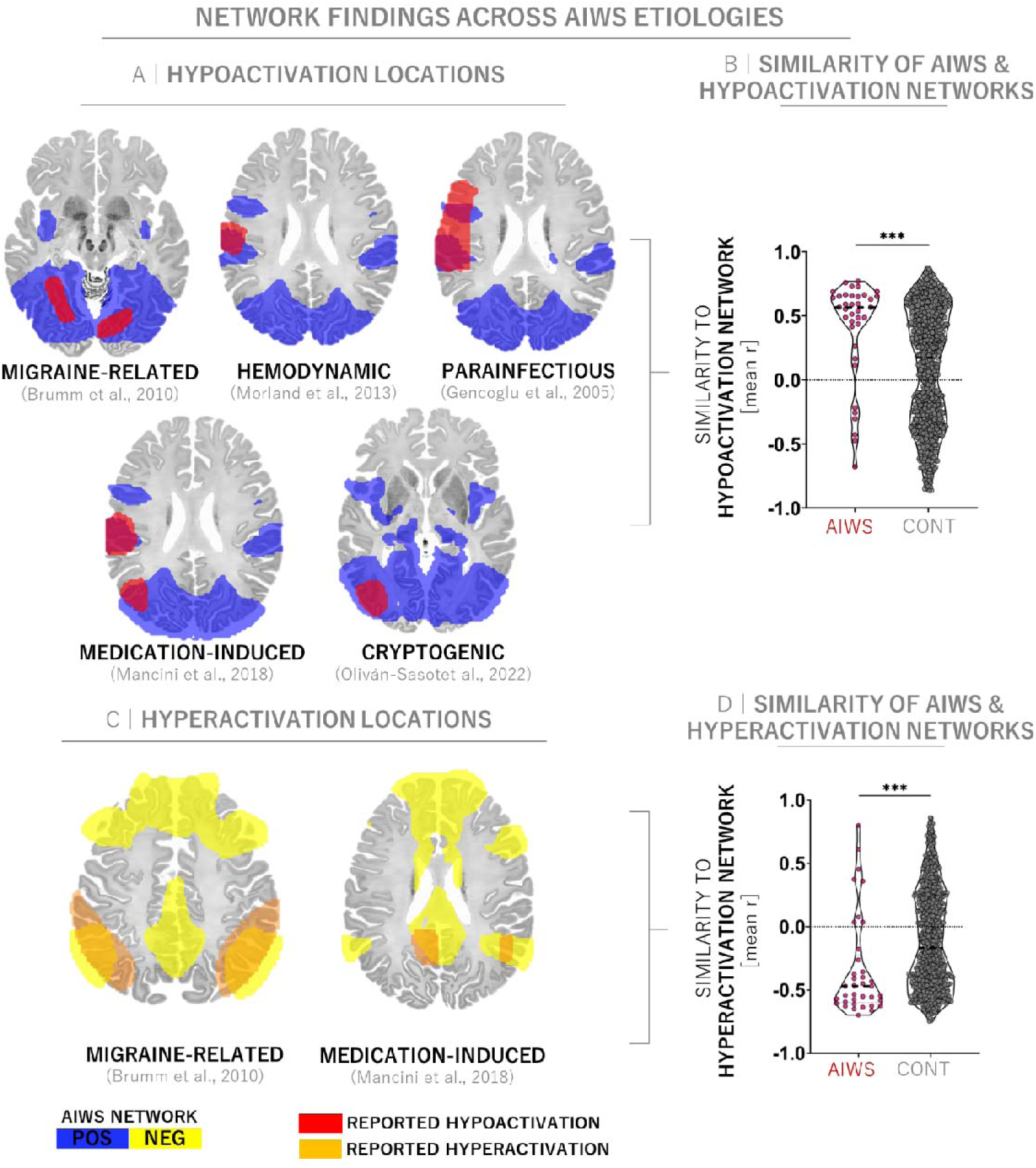
Alignment of AIWS-related neuroimaging findings across etiologies. A) Locations of brain hypoactivation observed during AIWS episodes (red) fall within our lesion-based AIWS network (blue, reproduced from Figure 4). B) Locations of hypoactivation showed a similar brain connectivity profile as lesions associated with AIWS (median r= 0.56, IQR 0.24), but not to as lesions associated with other neuropsychiatric syndromes as controls (CONT, p< .001, 95% CI [0.14, 0.44]). C) In contrast, reported regions of brain hyperactivation (orange) fell within a brain network with the opposite connectivity pattern of our lesion-based AIWS network (yellow). D) Locations of reported hyperactivations showed a brain connectivity pattern opposite to that of AIWS lesions, i. e. matching the “sign inverted” AIWS network (median r= −0.47, IQR 0.33) more than other neuropsychiatric lesion networks (p< .001, 95% CI [−0.11, −0.35]).

## Discussion

Lesions causing Alice in Wonderland Syndrome (AIWS), irrespective of subtype, are characterized by positive connectivity to the right extrastriate body area (EBA) as well as negative connectivity to the left inferior parietal lobule (IPL). This pattern of lesion connectivity was specific to AIWS when compared to over 1000 lesions causing other neuropsychiatric symptoms and aligned with neuroimaging findings from AIWS associated with various other etiologies. Overall, this suggests that despite clinical heterogeneity, AIWS is a coherent syndrome mapping to one specific brain network.

Our first key finding is that lesions causing AIWS are positively connected to the right EBA, a higher-order brain region preferentially activated by viewing bodies^36,37^. The EBA processes both visual and somatosensory information pertaining to bodies and body parts^36,44,45^. The right EBA is specifically involved in the perception of *one’s own body, others’* bodies^37,46^ and in discriminating *one’s own* and *others’* bodies^37,38,46,47^. While connectivity to a body-related brain region might be expected in AIWS-Self cases (which, by definition, always involve bodies), it may be surprising that this was also true for AIWS-Other cases (which do not necessarily involve perceptual distortions of bodies). Interestingly, the majority of the AIWS-Other lesion cases identified in our systematic search primarily involved bodies. This predominance is also apparent (but not highlighted) in prior studies of AIWS caused by other etiologies^3,5,48^. The specific connectivity of AIWS lesions to the EBA may help explain the high prevalence of body-related distortions across AIWS subtypes.

While connectivity to the EBA may help us understand why AIWS commonly involves bodies, it does not necessarily explain why AIWS invariably involves disruptions in size or scale. For example, patients could perceive too many bodies (as in polyopia) or a distortion of their color (as in cerebral dyschromatopsia)^49^. One possible explanation is that the phenomenology, or quality, of the perceptual distortion in AIWS may be determined by the disruption of a second brain region that is related to perception of size or scale.

Our second key finding is that the majority of AIWS lesions are also negatively connected to a region in the inferior parietal lobule, bordering the intraparietal sulcus. This area has been implicated in estimating magnitudes and dimensions, including object sizes, numerical distances^39,41,50^ and body dimensions^51^. Lesions to this brain region are linked to distortions in size and distance perception^52,53^, as well as disruptions in size constancy^53^ — a cognitive scaling mechanism that maintains constancy of an object’s size despite varying viewing distances^50^. This finding aligns with previous hypotheses suggesting disrupted size constancy in AIWS^54,55^ and may explain the frequent co-occurrence of size and distance distortions observed in AIWS cases.

Taken together, we hypothesize that lesions causing AIWS, due to their specific connectivity profile, simultaneously disrupt two higher-order brain functions related to body perception and judgement of size and scale. While the connectivity to the EBA may determine the predominant content of the perceptual distortion (i. e. body), connectivity to IPL may dictate its phenomenology (i. e. alterations in size and scale). These network-level findings may bridge previous theoretical frameworks implicating higher-order extrastriate visual areas and parietal multisensory areas in AIWS^3,5,8,54,56^. Similar theoretical frameworks have been implicated for lesions causing prosopagnosia^21^, delusional misidentifications^22^ and hallucinations^19^.

Finally, our findings from lesion-induced AIWS may lend insight in AIWS more generally. Our lesion-based network aligned with functional imaging in AIWS caused by other etiologies, including migraine, the most common cause of AIWS. Connectivity of area V3, which is implicated in the genesis of migraine aura^34,43^, exhibited a high similarity to lesions causing AIWS. This observed neuroanatomical convergence might provide a foundation to understand the occurrence of AIWS in up to 20% of patients with migraine^48^. Finally, the lesion-derived AIWS network aligned closely with neuroanatomically heterogeneous foci of altered brain activity during AIWS episodes from various other etiologies. This suggests that the network derived from focal brain lesion may lend insight into AIWS more generally, and that AIWS maps to the same neuroanatomical substrate independent of the etiology.

### Limitations

There are several limitations to our study. First, although our study is the largest study of AIWS lesion locations to date, it is retrospective and remains limited by a rather small number of lesion cases. Prospective neuroimaging studies in larger cohorts using targeted assessment of AIWS symptoms and comorbidities are needed to further validate our findings. Second, we lacked true three-dimensional images of the lesions, which limits voxel-wise lesion-symptom mapping techniques. Third, the lesion network mapping approach leverages a large functional connectivity dataset from healthy participants to approximate the connectivity of the lesion location in an “average” human brain. It is possible that individual functional connectivity of patients (prior to their lesion) could deviate from this average connection. It is also possible that our functional connectome lacked the spatial resolution to identify small clusters of lesion connectivity in areas like the brainstem. Finally, our results identify common hubs connected to lesion locations causing AIWS, but we are unable to determine if these hub regions themselves are dysfunctional in AIWS patients or how this dysfunction leads to AIWS symptoms.

## Conclusion

Lesion locations causing AIWS are defined by connectivity to two key regions previously implicated in body perception and size and scale judgements. These network-level findings have the potential to unify previous neuroanatomical and pathophysiological theories of AIWS and provide a mechanism by which a single lesion could simultaneously disrupt multiple cortical functions, leading to seemingly bizarre, but highly stereotypical perceptual alterations.

## Data Availability

Lesion masks in MNI space are available upon request to the corresponding author. The code for lesion connectivity analysis is freely distributed via the Lead-DBS toolbox (www.lead-dbs.org). The fully preprocessed version of the human resting-state fMRI-derived connectome is publicly available on Harvard Dataverse (https://dataverse.harvard.edu/dataset.xhtml?persistentId= doi:10.7910/DVN/ILXIKS and https://zenodo.org/records/4905738.

## Acknowledgements

The authors thank the scientific community involved in publishing the case studies underlying this work, Helen Friedrich for guidance on interpretation and visualization of anatomical data, and Ferdinand Friedrich for fruitful discussions.

## Data and code availability

Lesion masks in MNI space are available upon request to the corresponding author. The code for lesion connectivity analysis is freely distributed via the Lead-DBS toolbox (www.lead-dbs.org). The fully preprocessed version of the human resting-state fMRI connectome is publicly available on Harvard Dataverse (https://dataverse.harvard.edu/dataset.xhtml?persistentId= doi:10.7910/DVN/ILXIKS and https://zenodo.org/records/4905738.

## Author contact information

Maximilian U. Friedrich

Elijah C. Baughan

Isaiah Kletenik

Ellen Younger

Michael Ferguson

Charlie Weige Zhao

Calvin Howard

Amalie Chen

Daniel Zeller

Claudia Piervincenzi

Silvia Tommasin

Patrizia Pantano

Olaf Blanke

Sashank Prasad

Jared A. Nielsen

Michael D. Fox

