## Supplementary Information for "Lesions causing Alice in Wonderland Syndrome map to a common brain network linking body and size perception"

### Power analysis

Retrospective analyses are inherently limited by the achievable sample size. Effect sizes were approximated using three prior studies that contrasted lesion connectivity associated with presence versus absence of a perceptual distortion, resulting in values of 0.73^1^, 0.86^2^ and 2.2^3^. Assuming an α-error probability of .05 for an intended power of 0.85, the required sample sizes per group range from 10 to 66 lesion cases. This value falls well within previously reported cohort sizes of similar studies^4^. Power analysis was conducted in GPower (https://www.psychologie.hhu.de/arbeitsgruppen/allgemeine-psychologie-und-arbeitspsychologie/gpower).

### Syndromic definition

The characterization of AIWS in the literature has been heterogeneous^5–7^. Consistent with recent work^6,7^, we defined the syndrome as somatosensory and/ or visual distortions relating to the perception of size and scale of one’s own body (AIWS-Self) or distortions relating to the visual perception of size and scale of others’ bodies or objects in the environment (AIWS-Other). Cases with overlapping features were grouped as AIWS-Mixed. These distortions can include macropsia and micropsia (objects or body parts perceived as disproportionately large or small), telopsia and pelopsia (objects or body parts perceived as abnormally far or near) or hyper- and hyposchematia (external space or parts thereof perceived as abnormally large or small). These features are the most frequent and considered the defining core characteristics of AIWS across various etiologies^6,7^.

### Search strategies

The initial systematic literature search based on the “Preferred Reporting Items for Systematic Reviews and Meta-Analyses” (PRISMA) guidelines has been described in detail previously^8^. In brief, the key search terms were “macropsia” or “micropsia” or “macrosomatognosia” or “microsomatognosia” or “telopsia” or “pelopsia” or “porropsia” or “lilliputianism” or “aschematia” or “paraschematia” or “dysmetropsia.”

Our subsequent, independent PRISMA search contained as key terms "alice in wonderland" OR "macropsia" OR "micropsia" OR "macrosomatognosia" OR "microsomatognosia" OR "telopsia" OR "pelopsia" OR "body schema" OR "hyperschematia" OR "hyposchematia" OR "dysmetropsia" OR "’*metamorphopsia") AND ("lesion" or "stroke" or "haemorrhage" or "hemorrhage" or "inflamm*" or "plaque" or "tumor" or "trauma"). See Supplementary Figure 1 for full PRISMA flowchart.

We conducted a third search for studies reporting functional neuroimaging alterations during AIWS episodes. The key terms were: ("alice in wonderland" OR "micropsia" OR "macropsia" OR "macrosomatognosia" OR "microsomatognosia") AND ("functional imaging" OR "functional neuroimaging" OR "fMRI" OR "SPECT" OR "functional MRI"). This search yielded a total of 49 results. Of these, 5 studies documented locations of altered brain activity with sufficient quality for transfer into reference space, similar to lesion segmentation.

### Lesions associated with other neuropsychiatric disorders

We leveraged a large database of 1073 lesions across 25 neuropsychiatric disorders (Harvard lesion repository) from previous reports. These included: addiction/ addiction remission^9^, akinetic mutism and alien limb syndrome^2^, amnesia^10^, central visual disorders such as Anton syndrome, cortical blindness and blindsight^11,12^, aphasia^13^, asterixis and hemichorea-hemiballismus^14^, confabulation^15^, criminality^16^, delusional misidentification^17^, depression^18^ and mania^19^, focal epilepsy^20^, cervical dystonia^21^, hallucinations^1^, Holmes tremor^22^, disorders of consciousness^23^, parkinsonism^24^, post-stroke pain^25^, prosopagnosia^3^, cortical vertigo^26^.

| **Case ID** | **AIWS_Type** | **DOI / Pubmed identifier (PMID)** | **Foundational search** |
| --- | --- | --- | --- |
| AIWS_01 | A | 10.1186/s12883-020-01970-3 | Y |
| AIWS_02 | B | 10.21203/rs.3.rs-2090631/v1 | N |
| AIWS_03 | B | 10.21203/rs.3.rs-2090631/v1 | N |
| AIWS_04 | A | 10.1007/s00415-012-6827-5 | Y |
| AIWS_05 | B | 10.1371/journal.pone.0079938 | Y |
| AIWS_06 | A | 10.1016/j.neuropsychologia.2011.11.018 | N |
| AIWS_07 | B | 10.1007/s00415-007-0671-z | N |
| AIWS_08 | A | 10.1097/WNN.0b013e31826b70de | Y |
| AIWS_09 | A | 10.1097/WNN.0b013e31826b70de | Y |
| AIWS_10 | B | 10.1590/0004-282X20190094 | Y |
| AIWS_11 | C | 10.1080/13554794.2019.1656751 | Y |
| AIWS_12 | B | 10.1080/13554794.2018.1562079 | Y |
| AIWS_13 | B | 10.5692/clinicalneurol.cn-001081 | Y |
| AIWS_14 | B | 10.1016/j.nrl.2016.10.011 | Y |
| AIWS_15 | B | 10.4103/jnrp.jnrp_449_17 | Y |
| AIWS_16 | B | 10.1007/978-981-10-7668-8_52 | Y |
| AIWS_17 | B | 10.1007/s11682-015-9355-y | Y |
| AIWS_18 | B | 10.1155/2014/272084 | Y |
| AIWS_19 | B | 10.1016/j.nrl.2014.09.009 | Y |
| AIWS_20 | B | 10.1016/j.nrl.2014.09.009 | Y |
| AIWS_21 | B | 10.1016/j.ajem.2012.10.029 | Y |
| AIWS_22 | B | 10.1016/j.nrl.2010.07.029 | Y |
| AIWS_23 | B | 10.1016/j.jns.2010.05.015 | Y |
| AIWS_24 | B | 10.1016/j.jns.2010.05.015 | Y |
| AIWS_25 | B | 10.1136/jnnp.2006.100842 | Y |
| AIWS_26 | B | 10.1016/s0028-3932(99)00041-x | Y |
| AIWS_27 | B | 10.1093/brain/122.2.339 | Y |
| AIWS_28 | B | 10.1016/S0010-9452(08)70742-1N | Y |
| AIWS_29 | B | 10.1136/jnnp.57.1.73 | Y |
| AIWS_30 | B | 10.1136/jnnp.57.1.73 | Y |
| AIWS_31 | C | Kim YD, Ryu SY, Kim JS, Lee KS. Alice in Wonderland Syndrome in a Case with Infarct in the Right Medial Temporal Lobe. J Korean Neurol Assoc. 2006;24(4):364-366. | Y |
| AIWS_32 | B | PMID: 26111290 | Y |
| AIWS_33 | B | 10.1136/jnnp.54.1.68 | N |
| AIWS_34 | B | 10.2169/internalmedicine.8295-16 | N |
| AIWS_35 | B | 10.1136/jnnp.61.4.420-a | N |
| AIWS_36 | B | 10.1016/j.cortex.2020.02.012 | N |
| AIWS_37 | A | 10.1093/brain/awz179 | N |

*Supplementary Table 1 –* **Case overview.**

| **Lesion location** | **Right hemisphere (% cases)** | **Left hemisphere (% cases)** |
| --- | --- | --- |
| **Frontal cortex** | 0.0 | 8.1 |
| **Parietal cortex** | 5.4 | 5.4 |
| **Temporal cortex** | 27.0 | 8.1 |
| **Extrastriate occipital cortex** | 32.4 | 18.9 |
| **Striate occipital cortex** | 13.5 | 8.1 |
| **Cerebellum** | 0 | 5.4 |
| **Subcortical regions** | 2.7 | 2.7 |

*Supplementary Table 2* **Lesion topography of AIWS cases.** Note: Lesions that span more than one anatomical locations are counted for each location separately in this table. Therefore, the total percentages may exceed 100%.

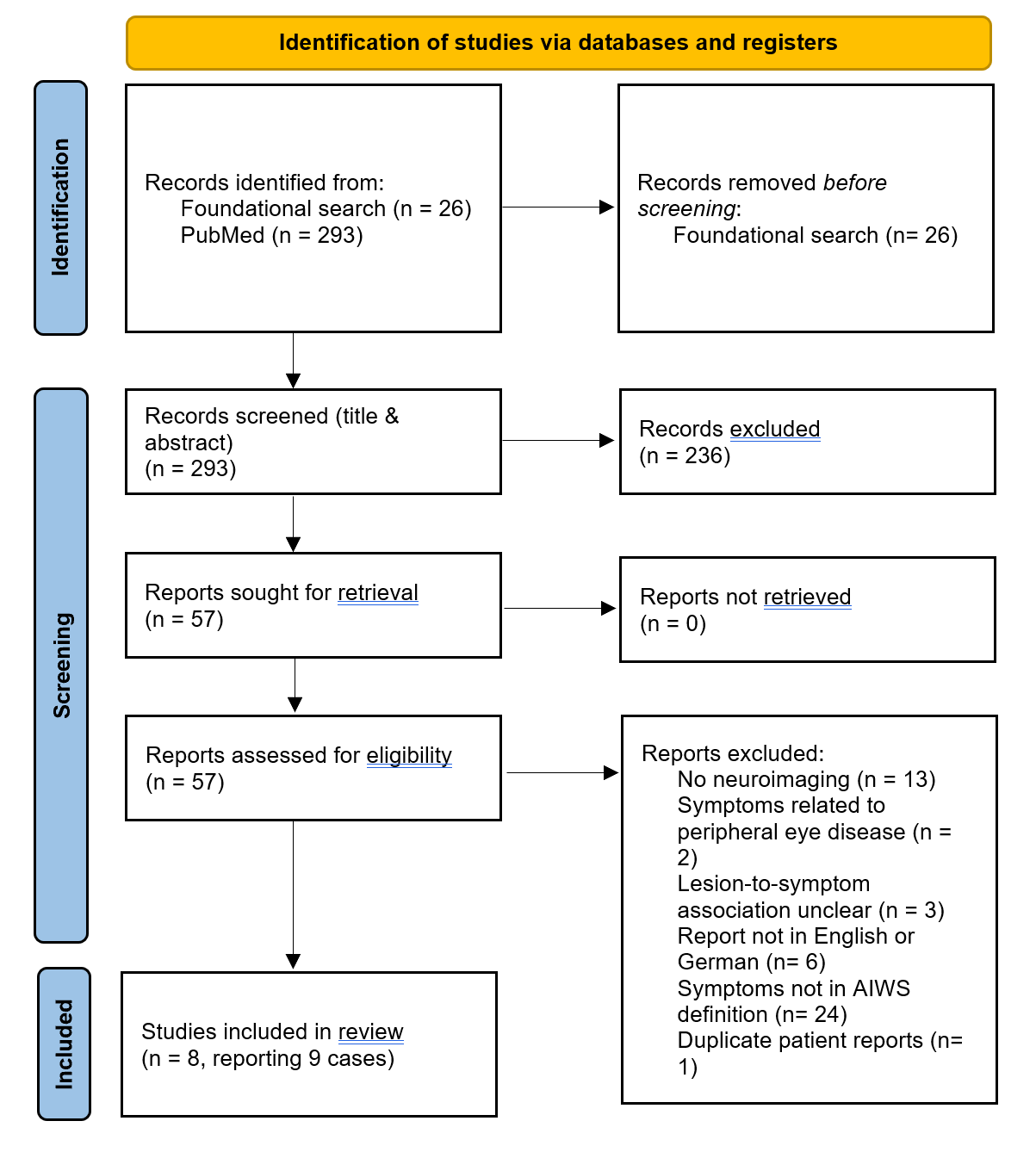

*Supplementary Figure 1.* PRISMA flowchart describing the search process to identify lesions associated with AIWS.

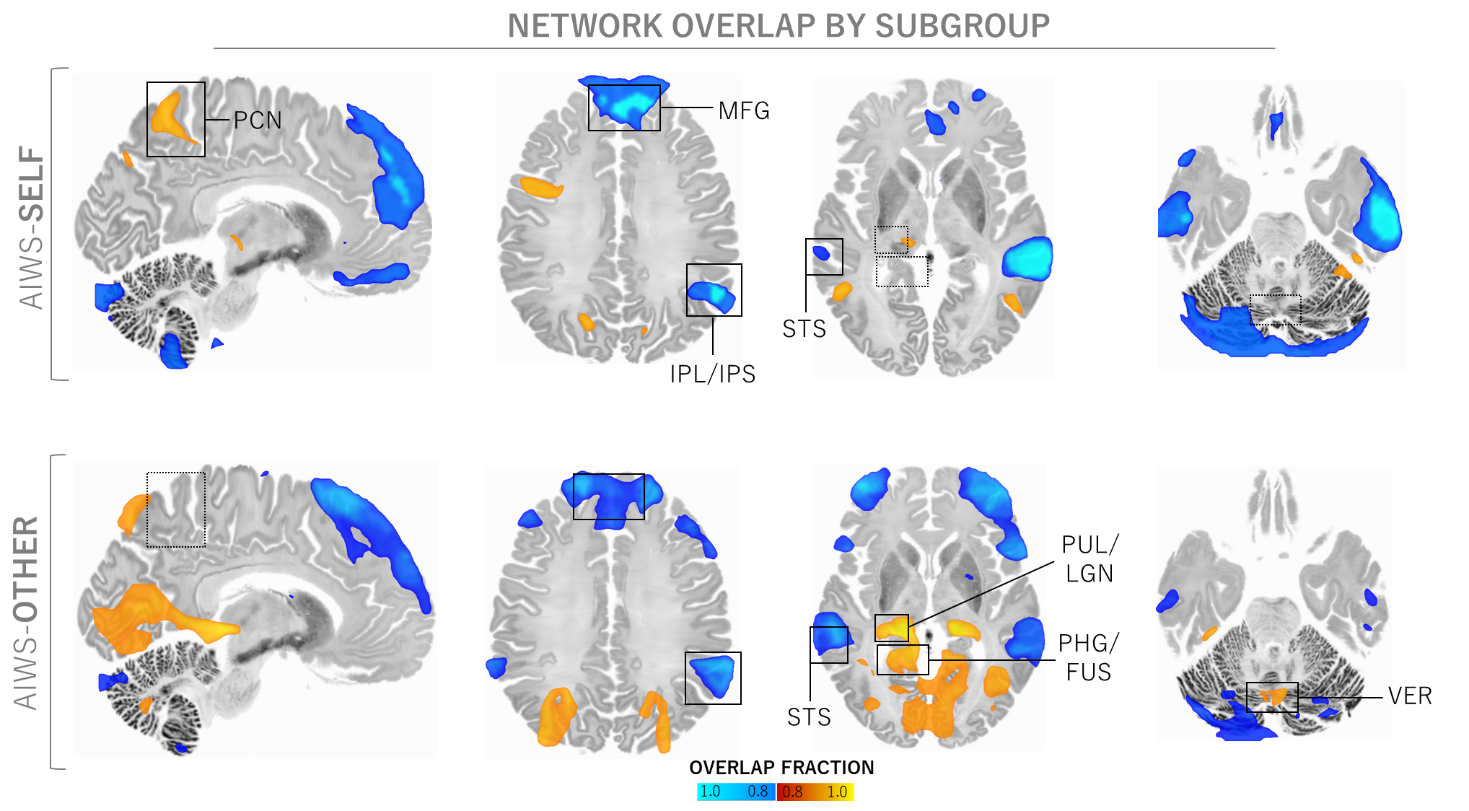

*Supplementary Figure 2 –* Lesion network overlap analysis, separated by AIWS clinical subtypes. Peak overlap regions of AIWS-Self include the precuneus (PCN), medial frontal gyrus (MFG), and inferior parietal lobule bordering the intraparietal sulcus (IPL/IPS). For AIWS-Other, regions of peak overlap are the MFG, IPL/IPS, the border zone of the parahippocampal and the fusiform gyrus (PHG/FUS), the superior temporal sulcus (STS) as well as the cerebellar vermis (VER). Warm colors represent positive lesion, cool colors negative lesion connectivity. Solid outlines highlight regions of shared overlap, dashed lines show regions with distinct connectivity between subtypes.

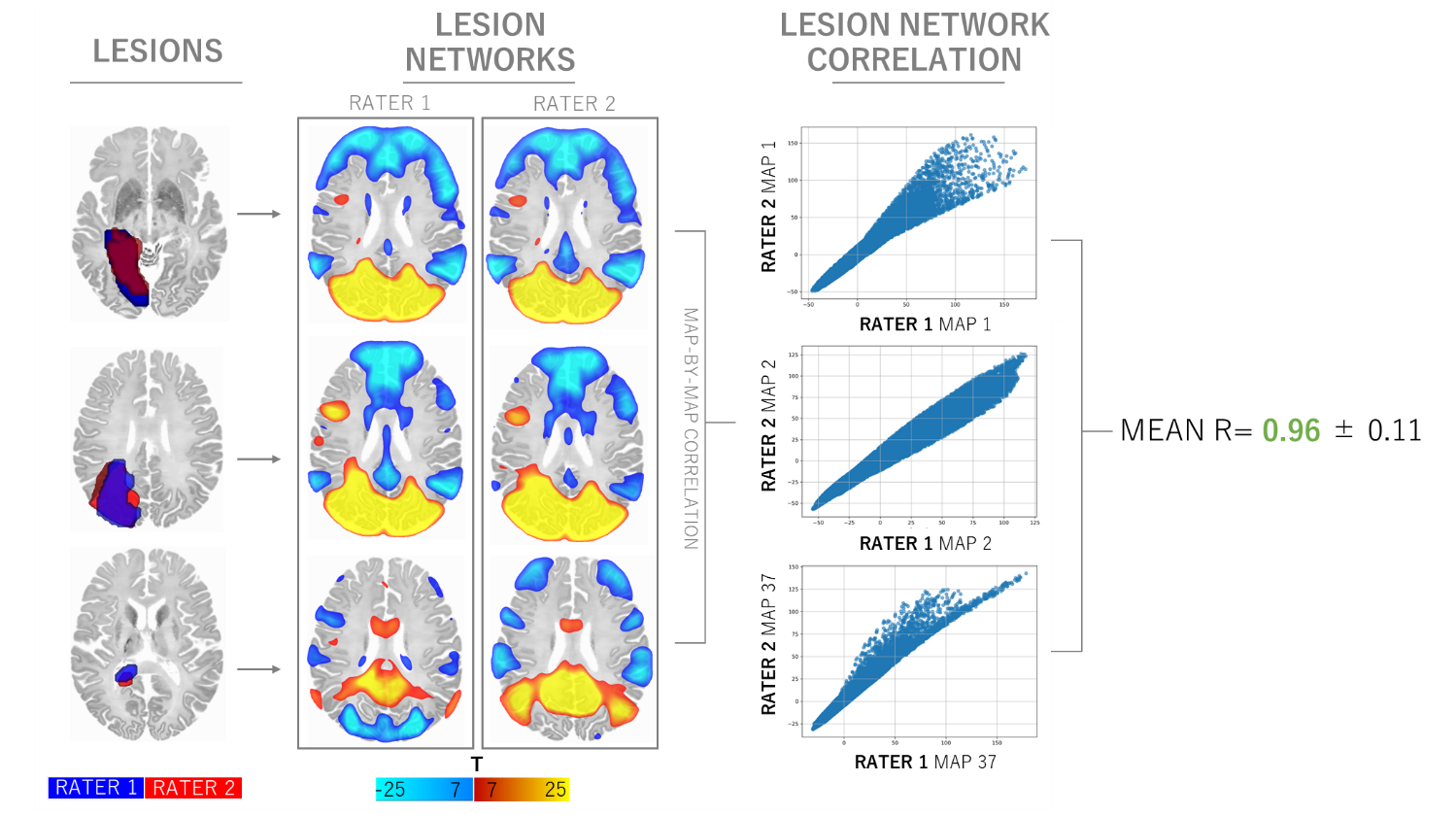

*Supplementary Figure 3* – **Lesion networks show consistency between two independent raters.** Lesions were independently traced by two raters (MUF, CWZ). Subsequent pair-wise correlations of the resulting lesion networks demonstrated a high level of concordance, with the resulting lesion networks being practically identical (mean Pearson r = 0.96 ± 0.11).

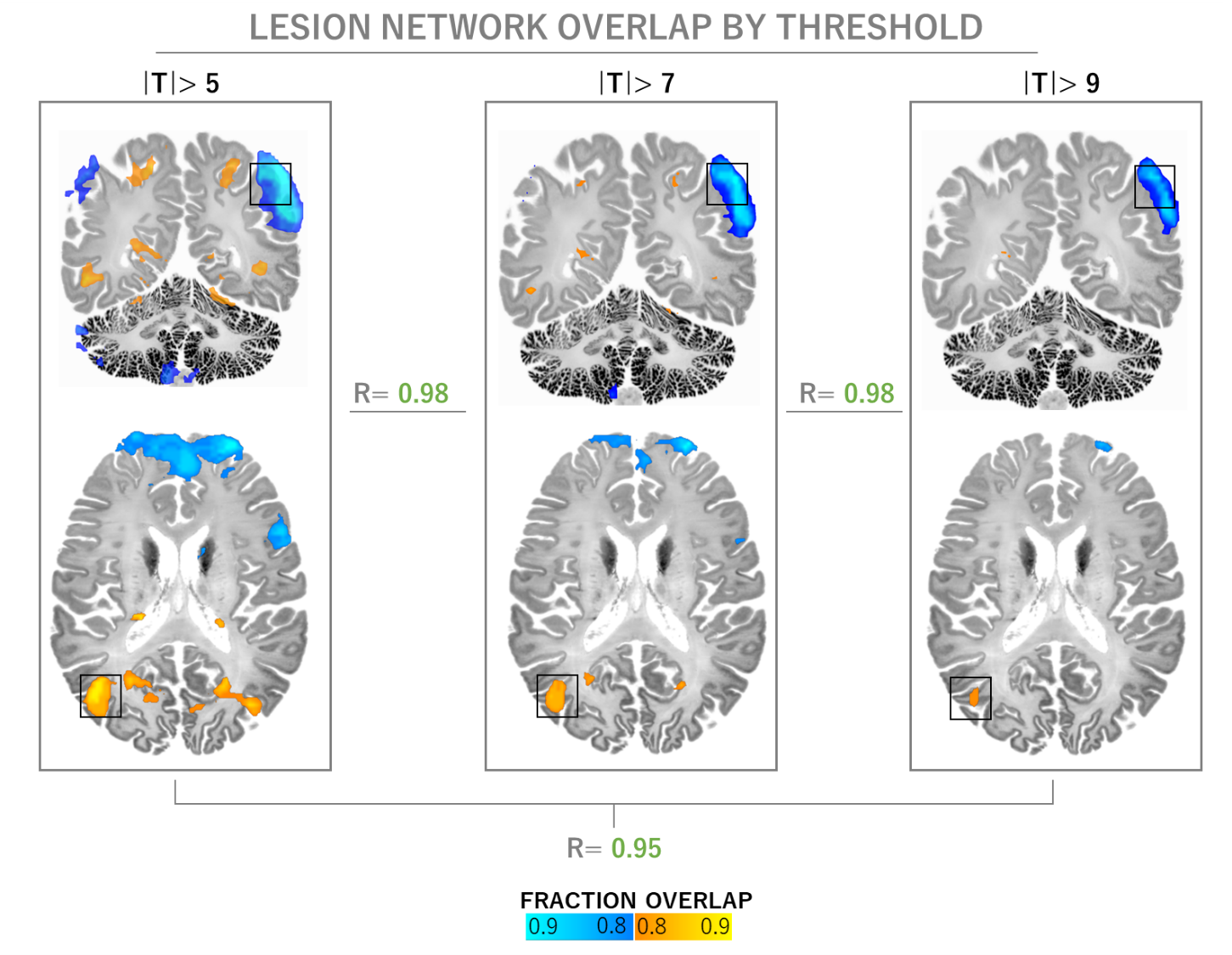

*Supplementary Figure 4 –* **Lesion network overlap maps are largely independent of statistical thresholding.** Fraction overlap map, masked for regions of at least 85% network overlap in both subtypes. This stability is substantiated by strong pair-wise Pearson correlations (r ≥ 0.95, p < .001), affirming that the identified connectivity patterns are robust and consistently replicated across various threshold levels. Notably, the key AIWS nodes in the right extrastriate body area (EBA) and the left inferior parietal lobule (IPL) consistently emerge in these maps, while the medial frontal gyrus finding does not survive more conservative thresholding. T= 5 corresponds to FWE-p< .05, T= 7 to FWE-p< 10^-6^ and T= 9 to FWE-p< 10^-9^.

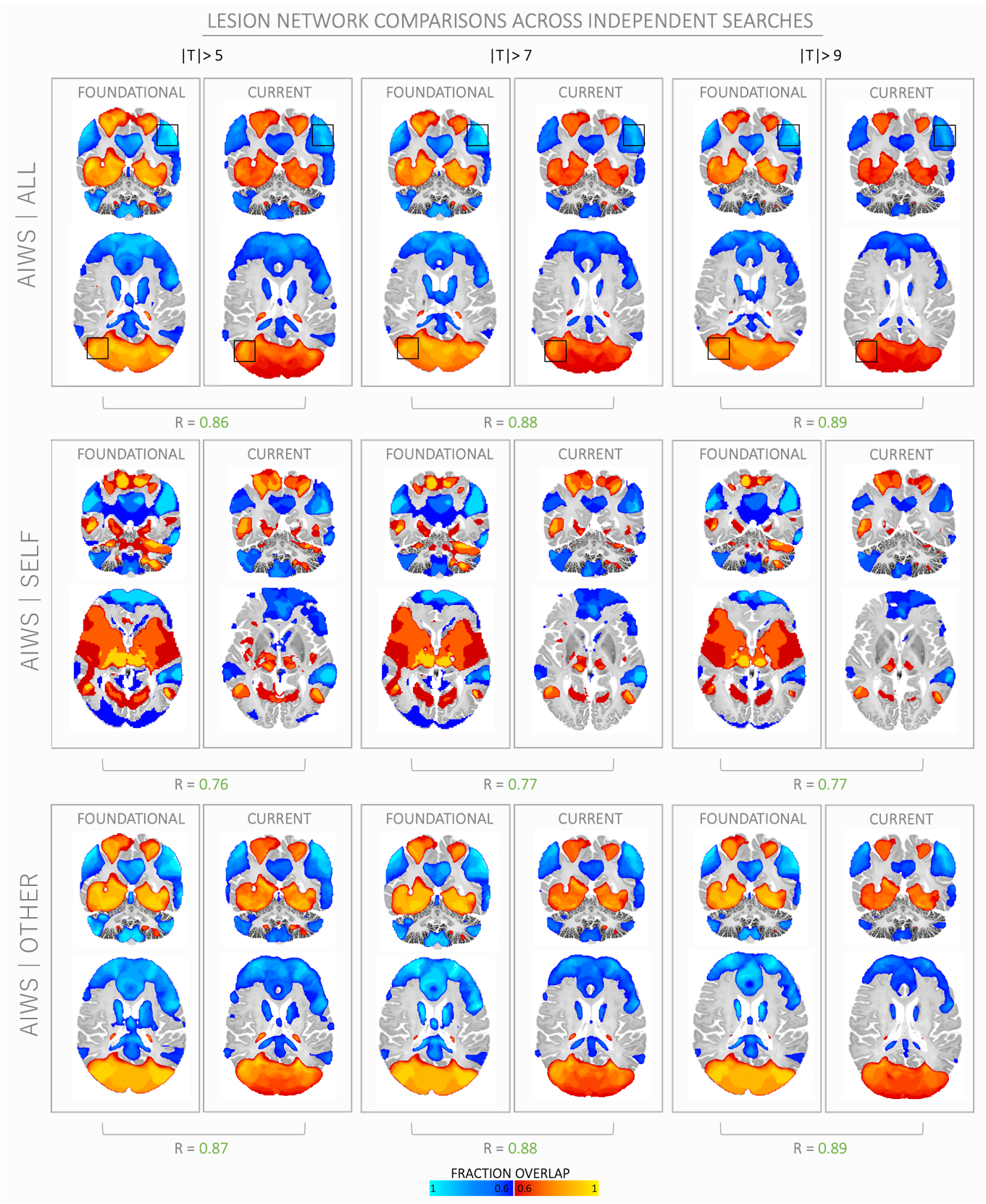

Supplementary Figure 5 – **Lesion networks are highly consistent across independent systematic searches and updated lesion inclusion criteria**. A foundational systematic search (conducted in November 2021 by CP) identified 30 lesions meeting inclusion criteria, which were then used in a lesion network overlap analysis. Results of this analysis are presented on the left for each threshold (|T| ≥ 5, 7, 9, left panels). A second, independent systematic search was conducted in September 2023 (MF), from which 37 lesions were included in the current lesion network analysis. Results of the current analysis shown on the right, for each threshold and broken down by AIWS subtypes. Despite the inclusion of additional lesions and independently conducted analyses, these lesion network overlaps demonstrate a robust pattern of functional connectivity underlying AIWS with peak connectivity areas localized in the right extrastriate cortex, left inferior parietal lobule as well as the medial frontal gyrus (map-by-map Pearson r= 0.76 – 0.89). Warm colors represent positive connectivity and cool colors represent negative connectivity. Lower overlap threshold chosen for a more comprehensive illustration of network topography.

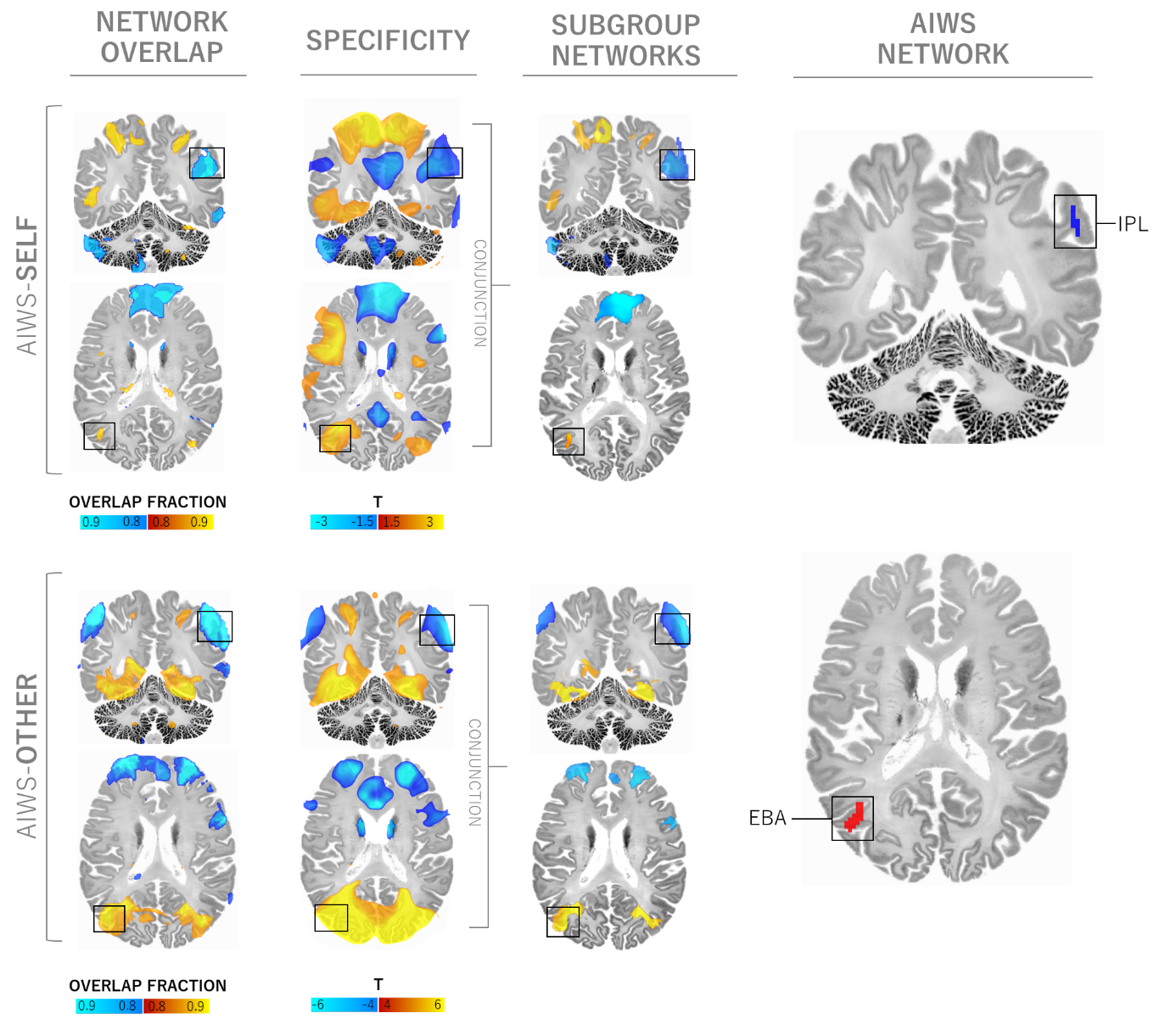

*Supplementary Figure 6.* **AIWS network findings are consistent across AIWS subtypes.** The conjunction analysis, repeated for each AIWS subtype separately, revealed a consistently shared connectivity pattern, notably involving the right extrastriate body area (EBA) and the left inferior parietal lobule (IPL). Although only the comparison of AIWS-Other versus neuropsychiatric controls reached voxel-wise significance (FWE-p< .05), the peaks observed in the unthresholded networks derived from the subtypes were closely aligned with the connectivity patterns previously identified in the overarching AIWS network. This alignment was particularly pronounced in the right EBA and at the border zone between the left IPL and the intraparietal sulcus.

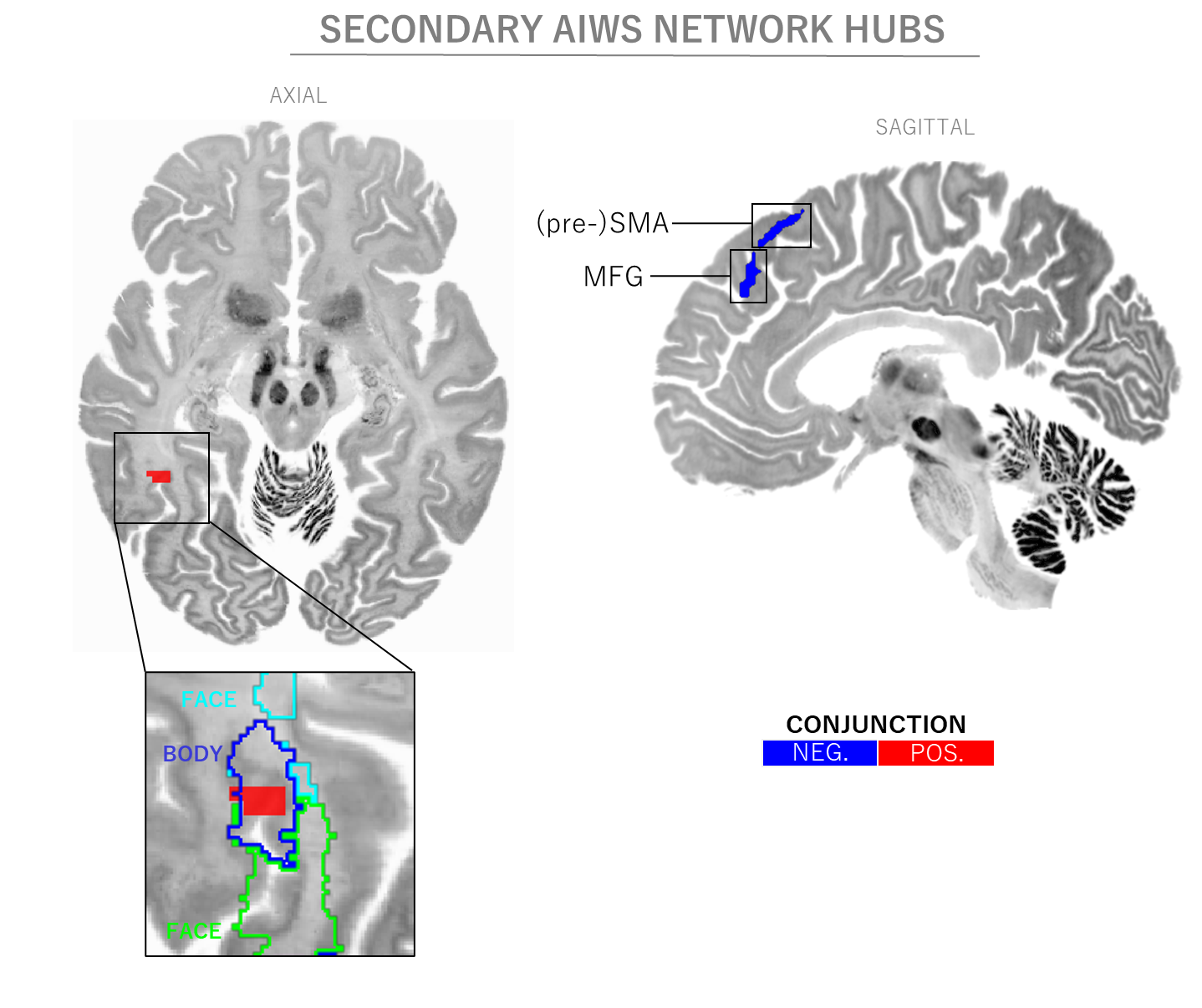

*Supplementary Figure 7 –* **Secondary connectivity findings.** In addition to the primary connectivity findings, the conjunction analysis revealed secondary, smaller clusters both sensitive and specific to AIWS. These clusters include the right superior parietal cortex (coordinates: 22, -72, 35; volume: 189mm³, not shown), left supplementary motor area (-5, 26, 57; 140mm³), right basal occipitotemporal (44, 47, -12; 84mm³), and left medial frontal cortex (-2, 40, 37; 75mm³). Notably, the right occipitotemporal cluster overlaps with a body-selective inferior temporal area, functionally similar to the extrastriate body area*^42^* (as shown in the inset).
